# Predicting Future Depressive Episodes from Resting-State fMRI with Generative Embedding

**DOI:** 10.1101/2022.11.17.22281138

**Authors:** Herman Galioulline, Stefan Frässle, Sam Harrison, Inês Pereira, Jakob Heinzle, Klaas Enno Stephan

## Abstract

After a first episode of major depressive disorder (MDD), there is substantial risk for a long-term remitting-relapsing course. Prevention and early interventions are thus critically important. Various studies have examined the feasibility of detecting at-risk individuals based on out-of-sample predictions about the future occurrence of depression. However, functional magnetic resonance imaging (MRI) has received very little attention for this purpose so far.

Here, we explored the utility of generative models (i.e. different dynamic causal models, DCMs) as well as functional connectivity (FC) for predicting future episodes of depression in never-depressed adults, using a large dataset (N=906) of task-free (“resting state”) fMRI data from the UK Biobank. Connectivity analyses were conducted using timeseries from pre-computed spatially independent components of different dimensionalities. Over a three year period, 50% of participants showed indications of at least one depressive episode, while the other 50% did not. Using nested cross-validation for training and a held-out test set (80/20 split), we systematically examined the combination of 8 connectivity feature sets and 17 classifiers. We found that a generative embedding procedure based on combining regression DCM (rDCM) with a support vector machine (SVM) enabled the best predictions, both on the training set (0.63 accuracy, 0.66 area under the curve, AUC) and the test set (0.62 accuracy, 0.64 AUC; p<0.001). However, on the test set, rDCM was only slightly superior to predictions based on FC (0.59 accuracy, 0.61 AUC). Interpreting model predictions based on SHAP (SHapley Additive exPlanations) values suggested that the most predictive connections were widely distributed and not confined to specific networks. Overall, our analyses suggest (i) ways of improving future fMRI-based generative embedding approaches for the early detection of individuals at-risk for depression and that (ii) achieving accuracies of clinical utility may require combination of fMRI with other data modalities.

## Introduction

Major depressive disorder (MDD) causes tremendous personal suffering and, amongst all medical conditions, has one of the highest burden of disease globally (GBD 2019 Mental Disorders Collaborators, 2022; Vos et al., 2020). It has a profoundly negative impact on social and occupational functions (Adler et al., 2006; Kupferberg et al., 2016) and is associated with increased risk for other mental and somatic (e.g. cardiovascular) disorders. After the onset of a first episode of MDD, there is a substantial risk for a long-term remitting-relapsing course (Eaton et al., 2008), accompanied by prolonged trial-and-error treatment attempts (Correll et al., 2017; Steffen et al., 2020). Prevention and early interventions are thus crucial for reducing the burden of MDD, both at an individual and societal level (Cuijpers et al., 2012, 2021). The challenge is to detect at-risk individuals early so that preventive measures and interventions can be administered in a timely and targeted fashion.

Detecting at-risk individuals requires prediction models that enable out-of-sample predictions about the future occurrence of (symptoms of) depression with clinically adequate accuracy. In the recent past, there have been numerous attempts to establish such models both in adolescents and adults, based on combinations of various data types, e.g. demographic, socioeconomic, cognitive, and clinical variables as well as motor activity (Caldirola et al., 2022; Chikersal et al., 2021; Gu et al., 2020; King et al., 2008; Librenza-Garcia et al., 2021; Lin et al., 2022; Na et al., 2020; Rocha et al., 2021; Rosellini et al., 2020; Sampson et al., 2021; van Eeden et al., 2021; Voorhees et al., 2008; Xu et al., 2019).

Neuroimaging has played a minor role in this endeavour so far. This may be partly due to difficulties of obtaining datasets that are longitudinal in nature and sufficiently large to allow for robust out-of-sample predictions. Several longitudinal magnetic resonance imaging (MRI) studies of depressive symptoms do exist (e.g. Barch et al., 2019; Pagliaccio et al., 2014; Papmeyer et al., 2016; Shapero et al., 2019), but almost all have small to moderate sample sizes and employ within-sample association analyses. However, association is not prediction: prediction requires out-of-sample analyses, i.e. “ … testing of the model on data separate from those used to estimate the model’s parameters” (Poldrack et al., 2020). A recent exception is the study by Toenders et al. (2021) which predicted depression onset out-of-sample, based on structural MRI (and other) data from a large sample of 544 adolescents. Concerning functional MRI (fMRI), however, we are aware of only one previous fMRI study (Hirshfeld-Becker et al., 2019) that has attempted out-of-sample predictions of future depressive episodes in hitherto depression-free individuals, albeit with a small sample (total N=33). The predictive value of fMRI for identifying individuals at risk for future depression is thus not well known.

One might wonder why fMRI should be considered at all for establishing predictor models of depressive episodes, given that fMRI data are more difficult to obtain and more costly than many other types of measurements? There are several reasons why fMRI – and particularly generative models for estimating connectivity – may have particular utility for clinical predictions. First, fMRI may afford high sensitivity since it assesses the functional status quo of neural circuits (Stephan et al., 2015), the biological level that is closest to psychiatric symptoms (Gordon, 2016). Second, clinical predictions are most valuable if they afford a mechanistic interpretation (Stephan et al., 2017); for example, this may guide the development of novel treatments. Analyses of functional interactions based on fMRI can potentially give insights into circuit mechanisms that increase risk for depression. Ideally, this requires generative models which offer an explanation how activity distributed throughout a circuit could have been generated (Stephan et al., 2015) and provide estimates of effective (directed) connectivity.

An approach that blends generative modeling with prediction is “generative embedding” (GE) (Brodersen et al., 2011, 2014; Frässle et al., 2020; Stephan et al., 2017). GE uses parameter estimates of a system (circuit) of interest, obtained by inverting a generative model, as features for subsequent machine learning (ML). This often improves prediction accuracy since the parameter estimates of a generative model offer a low-dimensional, de-noised representation of neural dynamics. Furthermore, provided the generative model is biologically plausible, GE may reveal which biological processes or properties (e.g. specific connections in a neural circuit) are most relevant for successful clinical predictions.

In this study, we used a large dataset (N=906) of task-free (“resting state”) fMRI data from the UK Biobank (Miller et al., 2016) to explore the utility of fMRI-based connectivity measures for predicting future episodes of depression in never-depressed adults. Over a three year follow-up period, half of the selected participants (N=453) exhibited at least one indicator of depression, according to clinical records and/or self-report, while the other half remained free from depression. Both groups were carefully matched with regard to 7 potentially confounding variables (age, sex, handedness, tobacco, alcohol, illicit drugs, cannabis).

We emphasise that the goal of this work was not to test whether predictions based on fMRI data are better or worse than predictions based on other data types, e.g. socioeconomic or clinical variables. Instead, because there are numerous options of utilising fMRI for predictive analyses, this initial study focused on fMRI only and assessed the relative performance of different connectivity approaches – including generative embedding based on different variants of dynamic causal modeling (DCM)^13^ as well as functional connectivity (FC) – for predicting future depressive episodes. Concretely, in our training set (N=724), we systematically combined different connectivity approaches with different ML classifiers, using nested cross-validation, and tested how well they predicted the occurrence of at least one indicator of a depressive episode over a follow-up period of three years. We then used the best-performing combination to make the same prediction in a held-out test set (N=182) that was completely independent from the training data. Notably, predicting the occurrence of indicators of depressive episodes represents a more challenging scenario than predicting a full clinical diagnosis of MDD. Our study can thus be seen as a “stress test” whether fMRI-based assessments of connectivity, and generative models in particular, are likely to be useful at all for early detection of at-risk individuals.

## Materials and Methods

The following sections describe the dataset and methodology used in this study. Briefly, the data consist of task-free fMRI measurements (i.e. unconstrained cognition or “resting state”) and questionnaire data from the UK Biobank (www.ukbiobank.ac.uk). Based on entries in UK Biobank, we selected participants that had good quality fMRI recordings and consistent questionnaire information that allowed us to assign them to one of two groups: a group that initially had no signs of depressive symptoms but exhibited indicators of depressive episodes (e.g. questionnaire data, prescription of antidepressants) within three years after the fMRI session (D+ group), or a control group that did not show any such indicators during the same period (D-group).

We used different connectivity metrics (different variants of DCM as well as functional connectivity, FC) in combination with different ML classifiers for prediction of future indicators of depressive episodes. DCM and FC analyses were applied to time series of rs-fMRI networks (with 6, 21, or 55 nodes) defined by independent components analysis (ICA) of the preprocessed “resting-state” fMRI (rs-fMRI) data and provided by UK Biobank. Posterior parameter estimates (DCM) and Pearson correlation coefficients (FC), respectively, served as input features to various discriminative classifiers. The classifiers were trained using nested cross-validation to avoid overfitting and to provide the best possible estimate of generalizability. Finally, the best models were chosen, and a prediction was made on held-out (and completely independent) test data.

It is worth noting that our analysis was pre-specified in an ex ante analysis plan, prior to performing any of the analyses. The analysis plan was time-stamped by uploading it to the Git repository of the Translational Neuromodeling Unit (TNU); it is available at https://gitlab.ethz.ch/tnu/analysis-plans/galioullineetal_ukbb_pred_depr. Furthermore, code reviews were performed by three of the co-authors (SF, SH and JH) who were not involved in the data analysis, both before the beginning of the analysis of the training data, and once again before running models on the test data. The code can be found at https://gitlab.ethz.ch/tnu/code/galioullineetal_ukbb_pred_depr.

### Dataset: groups with/without depressive episodes

The process of data extraction from the UK Biobank is summarised by Figure 1. To avoid confusion, it is worth explaining that participants of the neuroimaging branch of UK Biobank (which started in 2014) underwent two fMRI scans, approx. three years apart, each of which involved both task fMRI and rs-fMRI data. In this study, we only used the rs-fMRI data acquired during the first scan.

**Figure 1:**
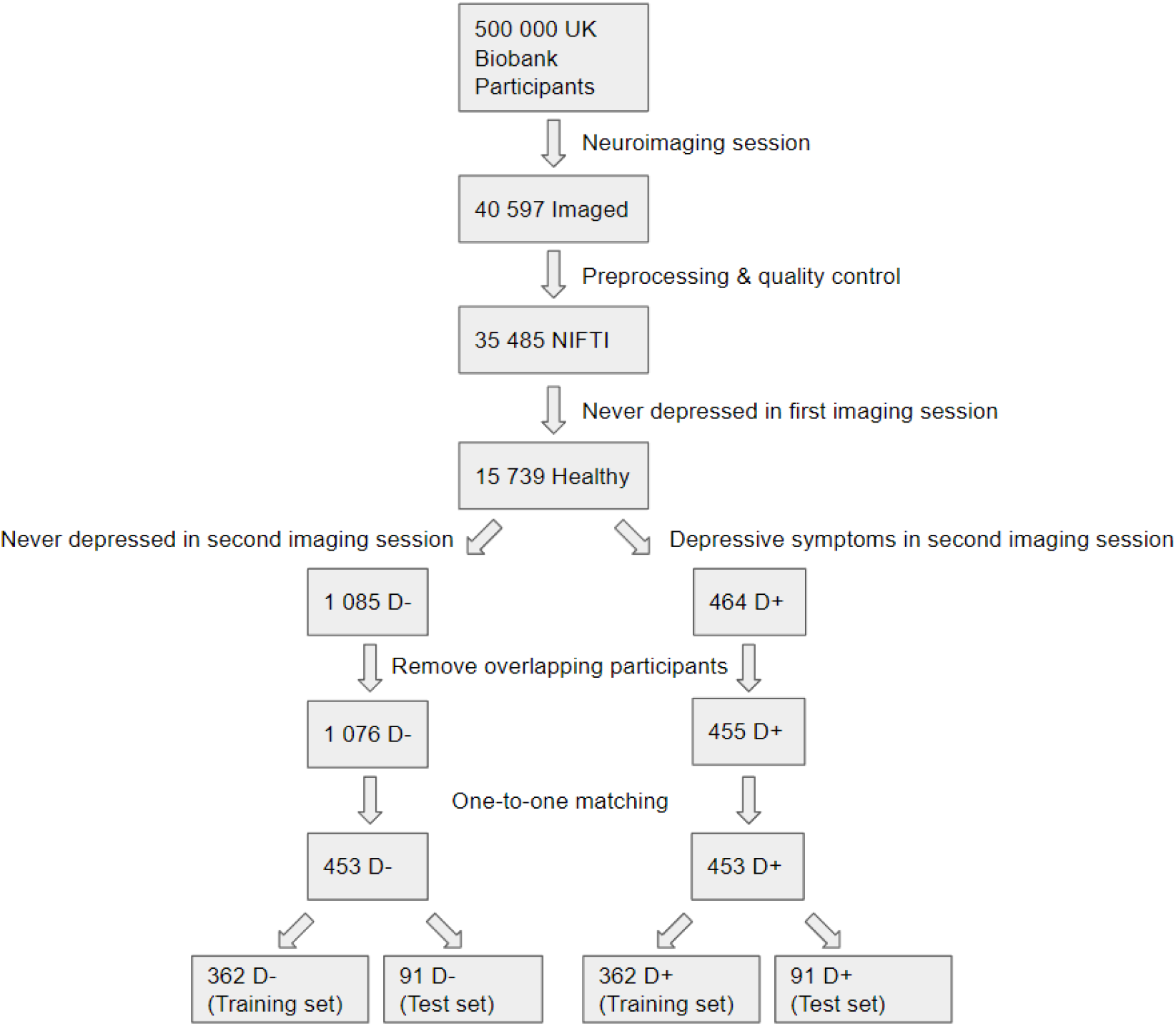
Flow diagram representing the dataset selection process. D-represents subjects who indicated no signs of depression, whereas D+ represents subjects who showed at least one indicator of depression.

Overall, selected individuals were required to have rs-fMRI data of good quality (as indicated by UK Biobank quality control) and no indication of any previous or current depressive episodes at the time of their first fMRI scan. From the subset of participants that fulfilled these criteria, we aimed to select two groups, one of which continued to indicate no signs of depressive episodes (D-group) three years after their first scan, and one that showed at least one indicator for at least one depressive episode over this three-year period (D+ group).

Concretely, we first identified participants who had both task (UKB field 20249-2.0) and “resting-state” (UKB field 20227-2.0) fMRI scans in NIFTI format, ensuring quality controlled images already preprocessed by UK Biobank (Alfaro-Almagro et al., 2018), resulting in 35,485 participants. In order to define the D-group, we chose the subset of participants who responded “no” to the questionnaire item “Looking back over your life, have you ever had a time when you were feeling depressed or down for at least a whole week?” (UKB field 4598-2.0) when they were first scanned (2014+). This resulted in 15,739 participants. We further shrunk this set by selecting those individuals who continued to show no evidence of depression in following years (2014 to 2019) and again replied, at the second fMRI session in 2019+, “no” to the previous question (UKB field 4598-3.0). This resulted in 1,085 potential D-participants who could be searched for matching criteria once the D+ group had been determined.

Concerning the D+ group, we also selected individuals from the set of 15 739 participants who had preprocessed imaging data and who – during their first imaging questionnaire (2014+) – indicated never having been depressed. Since the UK Biobank does not include information about the absence or presence of a clinical diagnosis of depression for all participants, we used multiple sources of information to identify indicators of depression. Specifically, we searched selected UK Biobank data fields which plausibly indicated the occurrence of at least one depressive episode in the years after the first fMRI scan. The following list summarizes the data fields in UKB and number of hits.

- Medical records in UKB:

- First Clinically Recorded Depressive Episode [UKB 130894] (5 hits)
- Clinical Depression-Related Encounter [UKB 41270] (31 hits)
- Prescription of Antidepressants [UKB 20003] (6 hits)
- Depression Diagnosis Report in UKB Assessment [UKB 20002] (12 hits)
- Self-report data in UKB:

- Depressed for at Least a Week Report [UKB 4598-2.0] (203 hits)
- Depression Diagnosis Report in Mental Health Questionnaire [UKB 20544] (90 hits)
- High Score (Coleman et al., 2020) on CIDI in Mental Health Questionnaire [UKB 20446] (165 hits)
- High Score (sum > 4) on Patient Health Questionnaire 3-subset [UKB 2050, 2060, 2080] (6 hits)

Overall, this resulted in 518 potential D+ participants. Since for any given participant a previous depressive episode could be reflected by multiple hits, we took the union of the above 8 sets of hits. This resulted in a total of 464 participants in the D+ group.

Having completed the initial definition of D+ and D-groups, we searched for data entries showing inconsistent or logically incompatible responses from participants (e.g. participants stating “never depressed for at least a week” but with a clinical report of depression). This process led to the removal of 9 participants in total, resulting in 455 participants in the D+ group and 1,076 participants in the D-group.

### Matching of participants and definition of training/test sets

To minimize any effects of potentially confounding variables, we matched participants with respect to multiple criteria. Specifically, for each D+ participant we tried to find a matching D-participant according to the following seven criteria (where a tolerance range was only allowed for age, as indicated):

- Sex (UKB field 31)
- Age ± 5 years (UKB field 34)
- Handedness (UKB field 1707)
- Tobacco smoking frequency (UKB field 1249)
- Alcohol consumption frequency (UKB field 1558)
- Ongoing addiction or dependence on illicit or recreational drugs (UKB field 20457)
- Historical cannabis consumption (UKB field 20453)

All but 57 D+ participants could be matched exactly. Out of these, 55 could be matched almost exactly, with at most one criterion deviating. Two D+ participants could not be matched and were excluded from further analyses. This provided us with a dataset of 906 participants in total: 453 D+ participants and 453 matched D-participants.

Finally, we performed an 80/20 split to partition the data into training and test sets. Both datasets were strictly separated from each other during data analysis to prevent any leakage of information that could affect the prediction results. We also addressed an unlikely, but theoretically possible, information leakage stemming from UK Biobank itself: the templates of major functional networks in the brain (Miller et al., 2016) which are offered by UK Biobank and which our study used for data extraction had been created using rs-fMRI data from the first 4,181 individuals in UKB. We resolved this potential problem by ensuring that all participants from this set that were also part of our extracted data were assigned to the training set. This resulted in patient/control training sets with 362 individuals each and test sets with 91 individuals (Figure 1).

### FMRI Data Analysis

Wherever possible we used data that is directly available on UK Biobank and did not require additional processing. The rs-fMRI data are of 6 minute duration (490 images, TR=0.735s), with a spatial resolution of 2.4mm isotropic, and were acquired with 8x multislice acceleration (Alfaro-Almagro et al., 2018). We used the data after the standard preprocessing pipeline executed by UK Biobank. The processing steps performed at UK Biobank included realignment, EPI distortion correction, and high-pass temporal filtering (with a 50s cut-off). The rs-fMRI data were further processed using single-subject spatial ICA decomposition using MELODIC in FSL (Jenkinson et al., 2012). The resulting independent components (ICs) were classified as signal vs. noise, and a cleaned version of the data was provided. UK Biobank then fed these data into a dual regression (Nickerson et al., 2017) based on a set of group-level templates of “resting-state” networks (based on data from 4’181 subjects) at dimensionalities of either 25 or 100. For subsequent analyses, 21/25 and 55/100 ICs were kept, as the others were found in previous work to be “…clearly identifiable as artefactual (i.e., not neuronally driven)” (Alfaro-Almagro et al., 2018).

ICs of resting-state data can be thought of as distinct functional networks (Smith et al., 2009), and interactions between these networks can be investigated by applying functional and effective connectivity methods to IC timeseries (for previous examples, see Goulden et al., 2014; Hyett et al., 2015; Motlaghian et al., 2022). In this work, we selected three sets of networks, which differed in the number of ICs included. Since we were interested in major functional networks implicated in depression (Brakowski et al., 2017; Kaiser et al., 2015), our first IC selection targeted the default mode network (DMN), central executive network (CEN), salience network (SN), and the dorsal attention network (DAN). The DMN and DAN are mapped (Miller et al., 2016) to IC indices 1 and 3, respectively, while the left/right SN and left/right CEN are mapped (Gratton et al., 2018; Shen et al., 2018) to IC indices 6, 5 and 13, 21, respectively. Furthermore, we considered IC sets of size 21 and 55 components (as provided by UK Biobank) in order to explore the impact of increasing the number of networks/ICs on prediction performance. The 55 components can be interrogated interactively via a web-based visualisation tool provided by UK Biobank: https://www.fmrib.ox.ac.uk/ukbiobank/group_means/rfMRI_ICA_d100_good_nodes.html

### Generative Embedding

Having completed the selection of timeseries, our analysis proceeded to generative embedding (GE). GE requires two choices: (i) a generative model, and (ii) a ML method that uses posterior estimates from the generative model as features.

Concerning the choice of generative models, our analysis considered three different variants of DCM that are suitable for task-free fMRI data: stochastic DCM (Li et al., 2011), spectral DCM (Friston et al., 2014), and regression DCM (Frässle, Harrison, et al., 2021). For all models and all IC sets, we assumed a fully connected network. As a reference, we also obtained functional connectivity estimates, based on Pearson correlation coefficients.

To invert stochastic DCMs, we used the *spm_dcm_estimate* function in SPM12, with a DCM struct as input which had its *Y*.*y* set to the 6 timeseries, ***a*** set to a 6×6 matrix of ones (fully connected network of endogenous connections), and *Y*.*dt* set to 0.735 (interscan interval). This resulted in a 6×6 matrix of effective connectivity estimates, giving us 36 features for subsequent ML. Due to its high computational complexity, it was not possible to run stochastic DCM with 21 and 55 IC timeseries.

For spectral DCM, we used the SPM12 function *spm_dcm_fmri_csd* with the same exact DCM struct as for stochastic DCM as input. The performance was notably faster than for stochastic DCM, but given that it still took a few hours to run on the Euler high-performance computing cluster of ETH Zurich and that the scaling of the computational complexity is supra-linear in the number of ICs (i.e., number of nodes in the DCM), we estimated that it would still take weeks or even months to run the entire analysis (i.e., inversion of the DCMs for all subjects) for 21 or 55 IC timeseries. Hence, just like in the stochastic DCM case, we restricted the spectral DCM analysis to 6 IC timeseries.

Concerning rDCM, its high computational efficiency enabled us to analyse networks consisting of more components (6, 21, and 55 ICs), resulting in 36, 441, and 3025 features, respectively. We used the rDCM code in TAPAS 4.0 (Frässle, Aponte, et al., 2021), with *Y*.*y* set to the respective time series, and *Y*.*dt* set to 0.735.

Finally, FC matrices were computed using the *corrcoef* function in MATLAB. Since these matrices are symmetric along the diagonal, and the diagonal is always 1, we took the upper triangle of these matrices to be our features, resulting in 15, 210, and 1485 features for the respective IC sets. It is important to note that FC does not capture any information about the directionality of connections, as opposed to the effective connectivity measures from the DCM variants described above.

### Classification

From the previous generative modeling, we had eight feature sets in place – functional connectivity for each IC set (6, 21, 55), stochastic and spectral DCM for 6 ICs each, and three rDCM feature sets for 6, 21 and 55 ICs. These feature sets were subsequently used as input to discriminative classifiers. Initially, we restricted all analyses to the training set data, and only touched the test data once we had selected a feature set / classifier combination that performed best. Regardless of the specific classifier chosen, the steps taken to arrive at reported metrics are the same.

Classifier training was performed using nested cross-validation (CV). Nested CV provides robustness against overfitting by optimizing hyperparameters in an inner CV loop while averaging the performance against other partitions of the data in an outer CV loop (Cawley & Talbot, 2010; Stone, 1974). In our case, we used 10 folds in the outer loop, and 5 folds in the inner loop. At the beginning of each iteration of the outer loop (before training with hyperparameter optimization), the confounds (sex, age, handedness, smoking, alcohol, illicit drugs, cannabis) were linearly regressed out using scikit-learn’s *LinearRegression* module. Then the data were normalized using the *StandardScaler* module and then the classifier was finally fit with the *GridSearchCV* module. This procedure yielded a set of performance measures for each feature/classifier combination (see Results).

After evaluation of the feature/classifier pairs on the training data, there are several possibilities how models fitted on the training data could be applied to the test data. First, we evaluated whether the feature set/classifier combination that had performed best on the training set generalised to the test set. Second, we performed a post hoc analysis in which we examined each feature set together with the classifier that had been optimal for this specific feature set on the training data.

In addition to evaluating classifiers based on their performance metrics, we also ran permutation tests to check for statistical significance of the classification results. These tests were run both on our training and test set. To generate an empirical null distribution for a given feature/classifier pair, we randomly permuted the labels while considering subject pairs between the D- and D+ groups, originating from the matching of confounds. This is done by identifying a pair and flipping their labels with 0.5 probability. For the resulting permuted labels, the classifier is trained again by re-running the entire nested CV procedure, yielding performance metrics under random conditions. This process is repeated many times (n = 1,000 in our case) to construct the empirical null distribution of performance metrics. We then compute the rank of the true performance metrics (obtained from the prediction without shuffling the labels) by calculating how many instances of the null distribution performed better. Dividing the rank by the number of permutations yields the p-value which we report.

A separate question concerned the choice of hyperparameters for the test set. While there are multiple options how hyperparameters for prediction on the test set could be chosen, we decided to use all data from the training set for optimising hyperparameters: we ran a non-nested 5-fold CV on the entire training set, picked the best-performing hyperparameters, and used those to predict on the test set. Other aspects relevant for classification on the test set, such as permutation testing, and regression of confounds were identical to the training set. Please see Figure 2 for a summary of the Materials and Methods described above.

**Figure 2:**
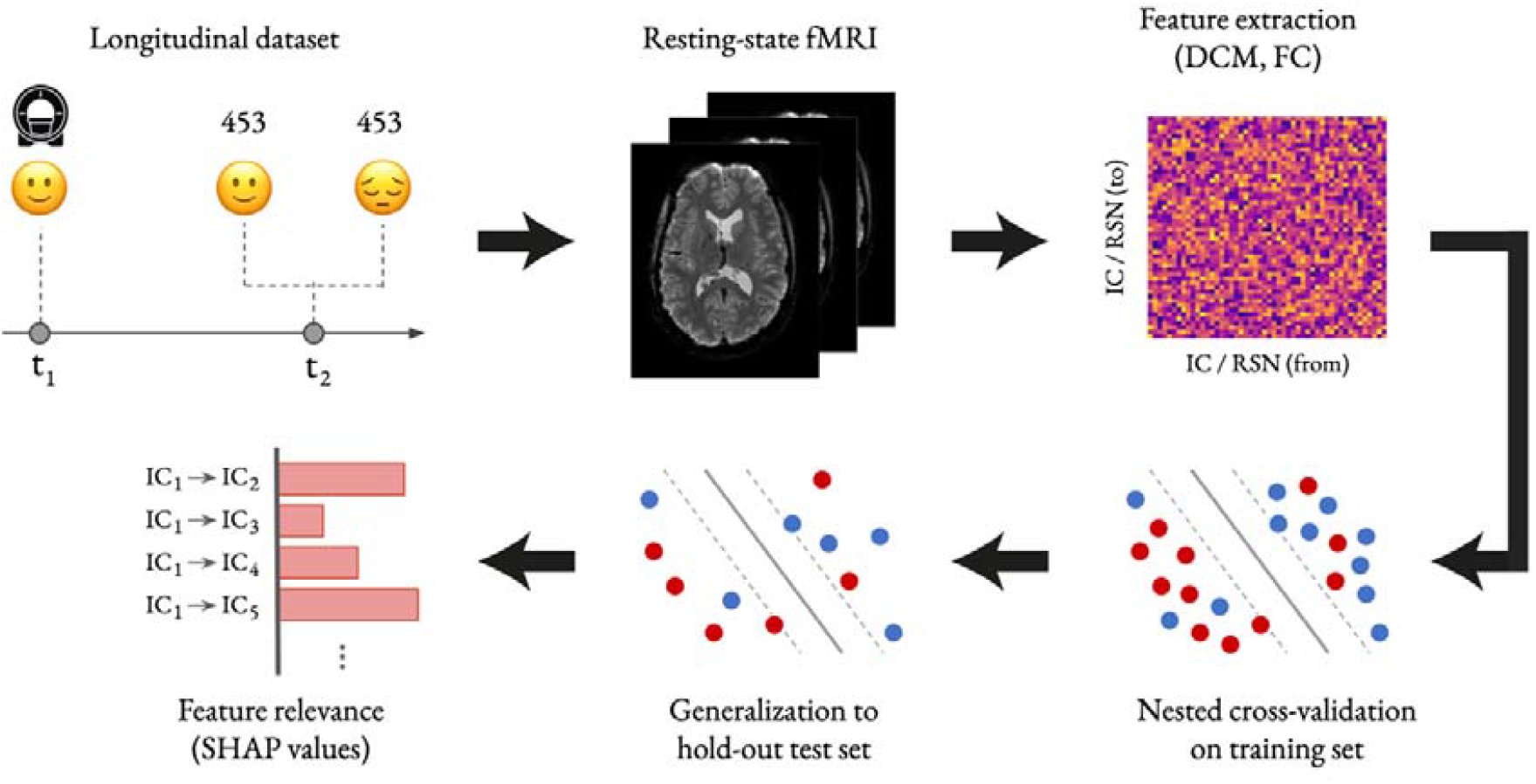
Illustration of the generative embedding pipeline utilized in the present study for predicting indicators of future depressive episodes. The pipeline comprises: Definition of the longitudinal dataset (top, left), identification of data features from the resting-state fMRI (rs-fMRI) data (top, middle), feature extraction, representing effective connectivity (dynamic causal modeling, DCM) or functional connectivity (FC) estimates amongst independent components (IC) or resting state networks (RSN) derived from the rs-fMRI data (top, right), nested cross-validation on the training set (bottom, right), generalization to the test set (bottom, middle), and feature relevance analysis based on SHAP values (bottom, left). Parts of the figure contain material from shutterstock.com (with permission).

Finally, we ran an interpretability analysis on our best-performing feature set/classifier combination (rDCM estimates based on 55 ICs and an SVM with a sigmoid kernel). This analysis based on SHAP (SHapley Additive exPlanations) (Lundberg & Lee, 2017), a generalisation of Shapley values from game theory (Shapley, 1953). For each feature, SHAP assigns an importance or attribution value that describes how much that feature contributes to the overall prediction. We used the *shap* software (https://github.com/slundberg/shap) to create a *KernelExplainer* that took as arguments:

- a sigmoid SVM classifier trained on rDCM with 55 ICs,
- a low-dimensional representation of the training data using *shap*.*kmeans* with five clusters (for computational tractability; see *shap* documentation)

Then, we computed the SHAP values using the explainer’s *shap_values* function which takes the test data as an argument. This gives us a SHAP value for each feature for each subject, which we process (mean of SHAP value magnitude across subjects) to obtain the average impact of each feature on model output magnitude.

### Choice and Implementation of Classifiers

A total of 17 classifiers were evaluated (please see Table 1 in the Results section), including six support vector machine (SVM) variants and three neural network (NN) variants. As described in the following, for most classifiers, we chose hyperparameters to optimize within the inner nested CV loop. For two classifiers (Gaussian naive Bayes and quadratic discriminant analysis) where hyperparameter tuning is less common, we kept scikit-learn’s default parameters.

**Table 1.**
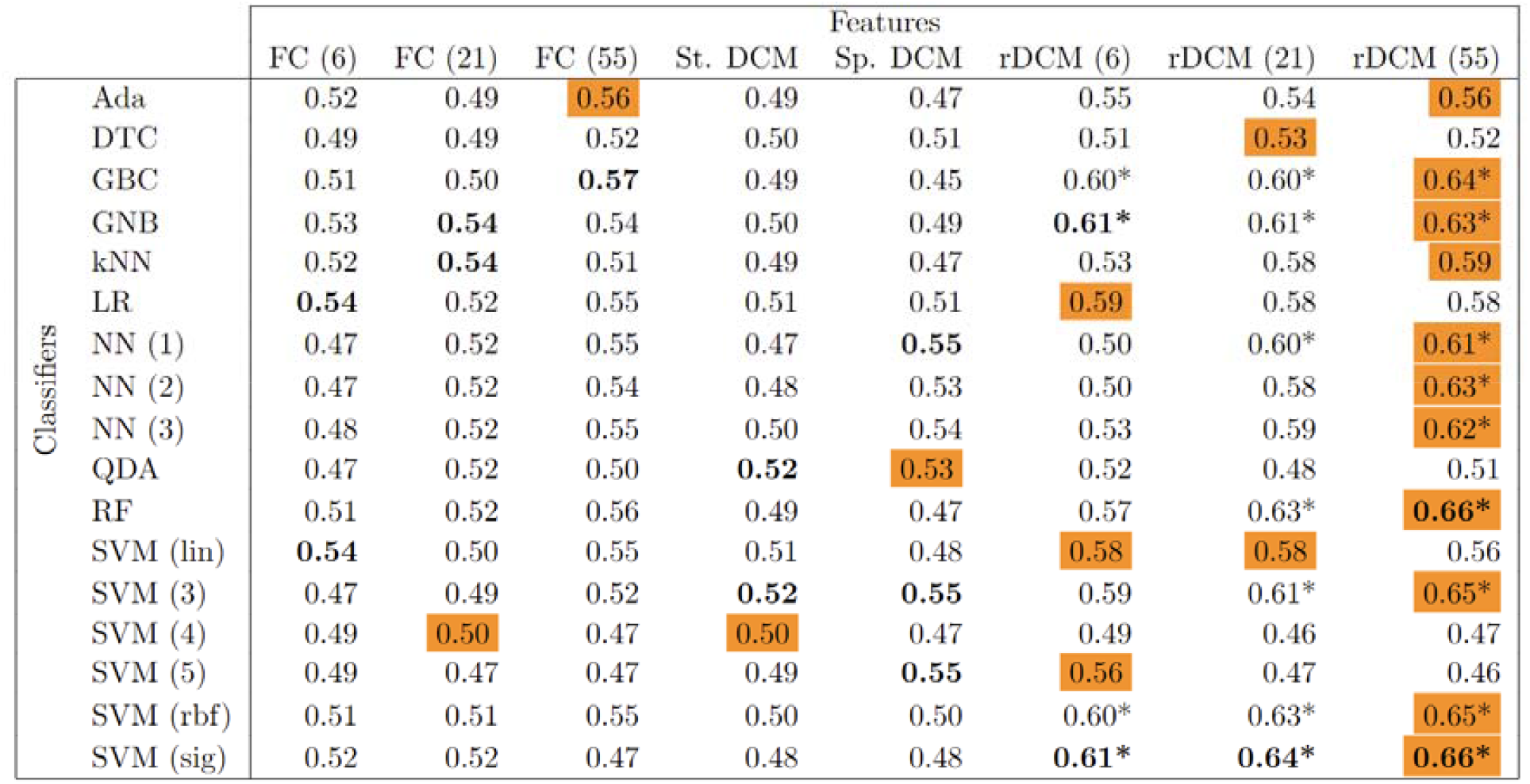
Summary table of AUROC for each feature/classifier combination as determined by nested cross-validation on the training set. Bold represents the best result across classifiers for a given feature set, orange shading represents the best result across feature sets for a given classifier, and a star denotes a statistically significant result (p ≤ 0.05). Ada: AdaBoost, DTC: Decision Tree Classifier, GBC: Gradient Boosting Classifier, GNB: Gaussian Naïve Bayes, kNN: k-Nearest Neighbors, SVM (lin): Support Vector Machine with linear kernel, LR: Logistic Regression, NN (n): Neural Network with n layers, SVM (n): Support Vector Machine with polynomial kernel order n, QDA: Quadratic Discriminant Analysis, SVM (rbf): Support Vector Machine with radial basis function kernel, RF: Random Forest, SVM (sig): Support Vector Machine with sigmoid kernel.

A first classifier was *logistic regression*. Following the default parameters of the scikit-learn version, we also used *L2* regularization, used *lbfgs* as our solver, and iterated at maximum 100 times. In the inner CV loop, we optimized for the regularization parameter C (0.01, 0.1, 1, 10, 100), which is the inverse of regularization strength (smaller values enforce stronger regularization).

For the *SVMs*, we made use of Platt scaling (Platt, 1999) to get probabilistic outputs for use in an AUROC (area under the receiver-operating characteristic curve) metric. We attempted classification with all types of kernels that scikit-learn has to offer, which include linear, radial basis function (RBF), sigmoid, and polynomial kernels with order 3, 4, and 5. We again treated the regularization parameter *C* (0.01, 0.1, 1) as a hyperparameter and additionally tuned gamma (1, 0.1, 0.01, 0.001) — the kernel coefficient for the RBF, sigmoid, and polynomial kernels.

We included three *neural network* variants (1, 2, and 3 hidden layers) which treat their layer sizes as hyperparameters. All other parameters are scikit-learn defaults (version 0.23.2), which means that – unlike logistic regression – the activation function used is actually ReLU (Rectified Linear Unit; Nair & Hinton, 2010).

Neural Network Hyperparameters

- 1 hidden layer sizes: 100, 150, 300, 500
- 2 hidden layers sizes: (100, 50), (150, 20), (300, 100), (500, 250)
- 3 hidden layers sizes: (100, 50, 5), (150, 20, 10), (500, 250, 50)

Ensemble methods combine multiple base models to (hopefully) produce better results than each individual model would have on its own. One such algorithm we used is *AdaBoost* (Freund & Schapire, 1997). We employed the scikit-learn default base classifier (decision tree) treating the number of estimators (30, 50, 70) as a hyperparameter. For a baseline comparison, we also attempted classification with a *single decision tree* with default scikit-learn parameters. Another ensemble method used in our classification is *gradient boosting* (Friedman, 2001) with 50, 100, 150 estimators as hyperparameters. Finally, we also tried *random forest* (Breiman, 2001), with options to tune 100, 500, 1000 estimators. For random forest, we additionally treated the maximum tree depth (10, 30, 60) as a hyperparameter.

We also explored prediction with three supervised learning algorithms that do not fall under the previous categories. Namely, *Gaussian naive Bayes* (Zhang, 2004), *quadratic discriminant analysis* (Cover, 1965) – both of which use scikit-learn default parameters – and *k-nearest neighbors* (Cover & Hart, 1967) where we treated the number of neighbors (3, 5, 7, 9) and leaf size (20, 30, 40) as hyperparameters.

We used a variety of metrics to evaluate classifier performance in order to ensure a holistic view and to avoid potential pitfalls (such as overemphasizing the importance of one metric). We report recall (sensitivity), precision (positive predictive value), F_1_ score, accuracy, and AUROC (area under the receiver-operating characteristic curve).

### Deviations from the original analysis plan

Our analyses were pre-specified and are described in a time-stamped analysis plan (https://gitlab.ethz.ch/tnu/analysis-plans/galioullineetal_ukbb_pred_depr). We subsequently extended this analysis plan in three ways:

1. We extended the coverage of networks and, in addition to the 6 networks (represented by IC timeseries), also considered sets of networks consisting of 21 and 55 ICs, as provided by UK Biobank.
2. We extended the connectivity methods by considering functional connectivity (Pearson correlation coefficients) in addition to variants of DCM as generative models.
3. In addition to SVMs, we decided to test a larger set of classifiers in order to avoid that our results may depend on the particular choice of classifier.
4. We included an analysis of feature importance on the test set using SHAP values.

The decision to extend the analyses in this manner took place before any prediction analyses of the training or test data were conducted.

## Results

We first present the performance of the cross-validated classifiers for the training dataset and then proceed with the most promising feature/classifier combinations to the test dataset. Note: since our dataset is balanced, accuracy as a metric implies balanced accuracy.

### Training set

Table 1 provides an overview of prediction performance on the training set in the nested cross-validation setting. Altogether, 17 different classifiers were evaluated (including four SVM variants and three neural network variants). We report AUROC of all the features run with each classifier (Table 1) and we also report all metrics for the five best feature/classifier combinations (Table 2).

**Table 2.**
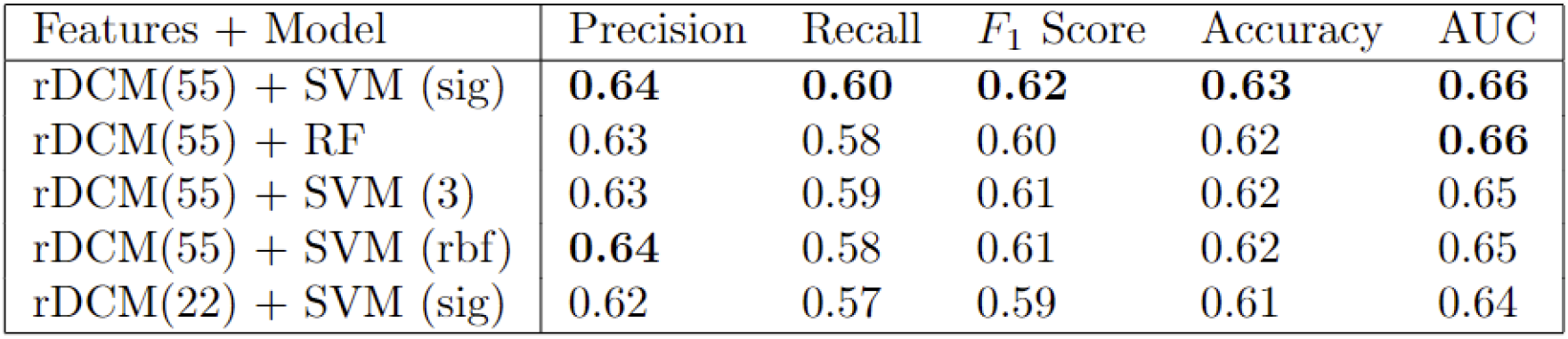
Summary of all metrics on the top five feature set/classifier combination as determined by nested cross-validation on the training set. Bold indicates the best model for the given metric.

Having run all feature set/classifier pairs on the training data using nested cross-validation, we found that a sigmoid SVM paired with an rDCM taking 55 ICs – referred to subsequently as rDCM(55) – as input performed best (Tables 1, 2). In terms of performance, applying a sigmoid SVM to rDCM connectivity estimates based on 55 ICs resulted in an AUROC of 0.66 (Figure 3A). The other performance metrics for this combination were: precision=0.64, recall=0.60, F1 score=0.62, accuracy=0.63 (Table 2).

**Figure 3:**
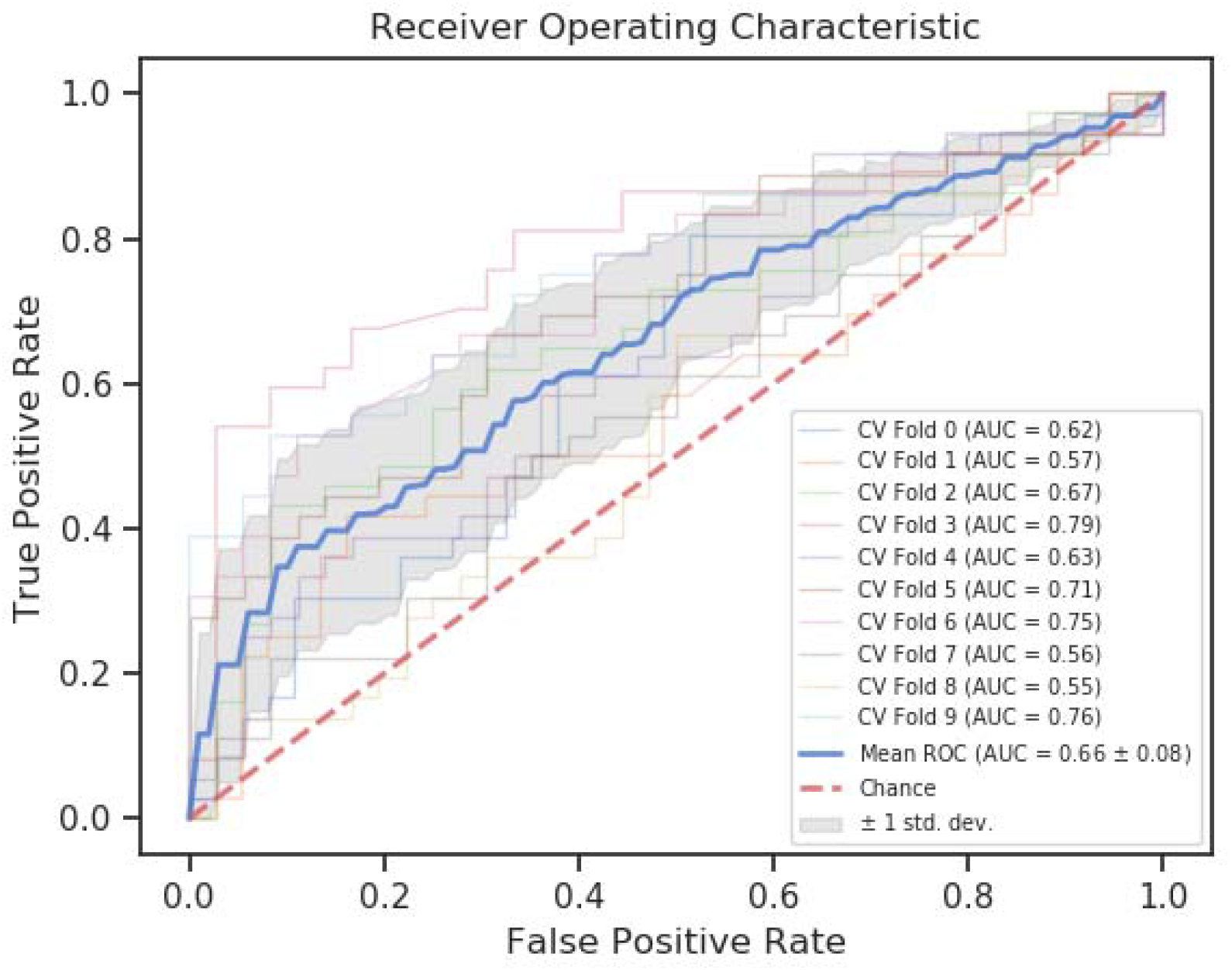
ROC curve for sigmoid SVM paired with rDCM(55) on the training set run with nested cross validation.

In general, connectivity estimates by rDCM enabled better predictions, regardless of classifier (see Table 1, orange shading): for 15 out of the 17 classifiers tested, one of the rDCM feature sets resulted in the best AUROC (in 11/15 cases, the best feature set was rDCM(55)). Furthermore, SVMs tended to perform better than other classifiers, with SVM variants having highest accuracy for 6 out of 8 connectivity feature sets. In particular, the sigmoid SVM was the best-performing classifier for 3 feature sets, more than any other classifier.

Based on these results, we chose rDCM(55) with sigmoid SVM to move forward to the test set. The test data had not been touched up until this point to prevent any leakage of information and ensure a thorough verification of the generalizability of our prediction model.

### Test set

The prediction of the best-performing approach on the training set generalized to the test data: the application of a sigmoid SVM to connectivity estimates by rDCM (55 ICs) from the test set showed an AUROC of 0.64 (Figure 4A). This prediction performance was significantly above chance: Figure 4B shows that the achieved accuracy of 62% is well outside the null distribution generated by predictions on randomly permuted labels (p<0.001).

**Figure 4:**
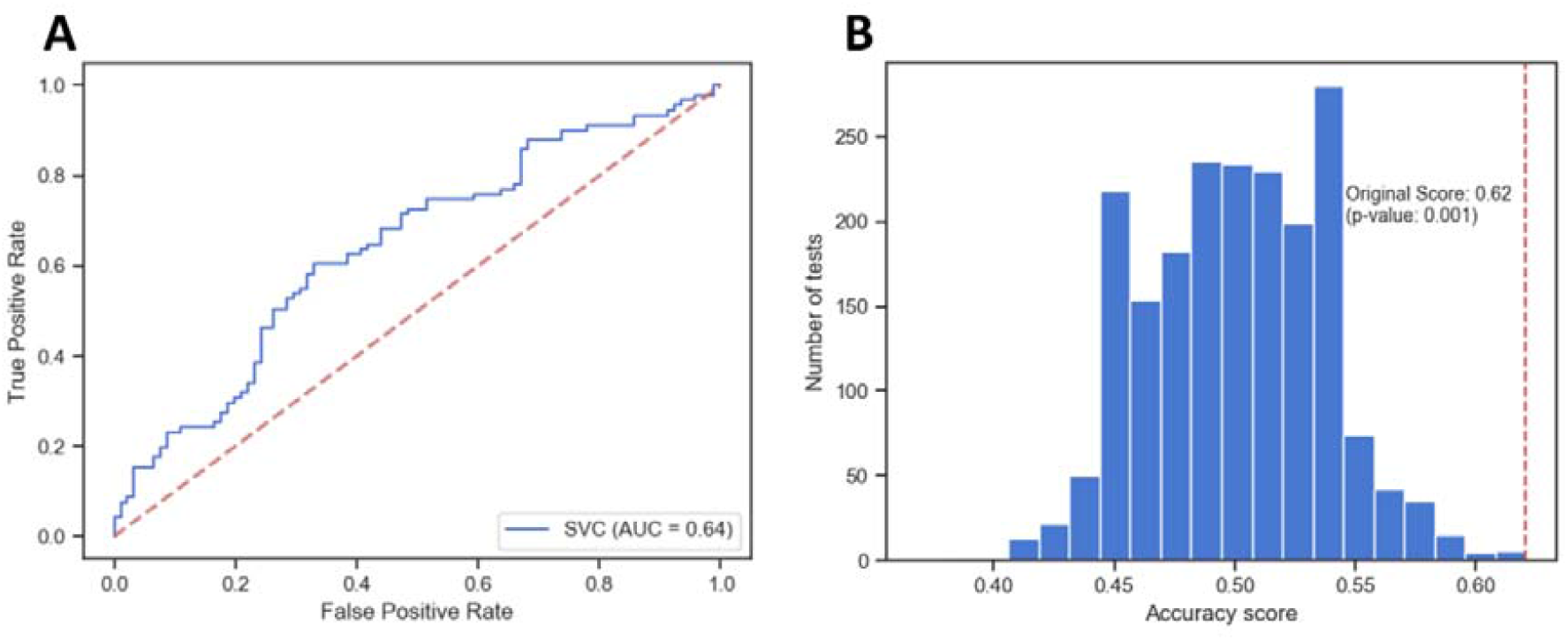
(A) ROC curve of rDCM (55 ICs) with sigmoid SVM run on test data. (B) Permutation test (n=1,000) run on test data with accuracy as the metric.

To get a better understanding of the generalization performance we conducted a post-hoc analysis on other well-performing feature/classifier pairs (Table 3) from the nested cross-validation and assessed their performance on the test data. We defined “well-performing” as the best classifier in general, but for feature sets where another classifier performed better, we selected the latter instead. From the 13 classifiers tested post-hoc, only three other pairs had above-chance performance on the test set, namely rDCM (21 ICs) with sigmoid SVM (58% accuracy, p=0.019), functional connectivity (6 ICs) with sigmoid SVM (59% accuracy, p=0.007), and functional connectivity (21 ICs) with Gaussian Naïve Bayes (59% accuracy, p-value=0.012). All of these performed worse with at least a 3% drop in accuracy, leaving the rDCM (55 ICs) with sigmoid SVM as the best-performing model overall on the test data.

**Table 3.** Summary of all metrics on the top five feature/classifier combinations on the test set.

| Features + Model | Precision | Recall | $F_1$ Score | Accuracy | AUC |
| --- | --- | --- | --- | --- | --- |
| rDCM(55) + SVM (sig) | **0.63** | 0.56 | **0.60** | **0.62** | **0.64** |
| FC(21) + GNB | 0.59 | **0.60** | 0.59 | 0.59 | 0.58 |
| FC(6) + SVM (sig) | 0.59 | 0.59 | 0.59 | 0.59 | 0.61 |
| rDCM(21) + SVM (sig) | 0.60 | 0.51 | 0.55 | 0.58 | 0.59 |
| FC(6) + Log Res | 0.57 | 0.52 | 0.54 | 0.57 | 0.61 |

Finally, we computed the SHAP values (Figure 5) for our best-performing model, the sigmoid SVM classifier paired with rDCM(55). This assesses the contribution of each connection to the prediction performance. Since rDCM provides estimates of effective (directed) connectivity, we have two SHAP value estimates for each IC, one for the outgoing connection, and one for the incoming connection. We visualized the top 100 SHAP values as a circular plot, where each IC is shown twice, and the bottom half represents the associated values for the outgoing connections. The width of each displayed connection reflects the magnitude of the SHAP value, and the width of the coloured IC label on the circle represents the cumulative SHAP value for outgoing or incoming connections of that node. Figure 5 shows that connections with the top 100 SHAP values were not confined to a few networks but included almost all ICs, with very few exceptions. Put simply, during the “resting” state of unconstrained cognition the participants were in, the most predictive connections were found all over the brain.

**Figure 5:**
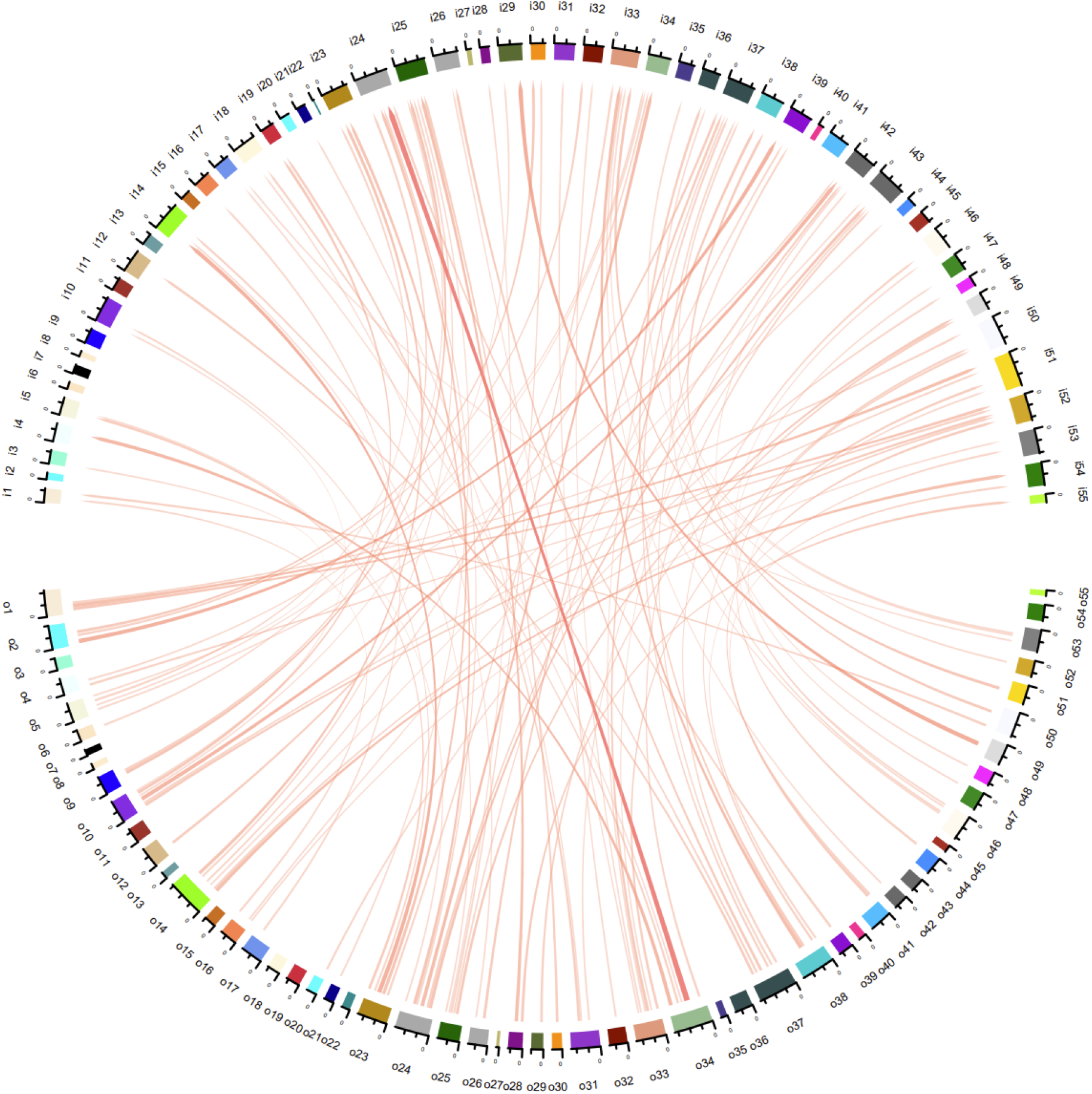
Top 100 Shapley values for sigmoid SVM paired with rDCM(55) on the test data. Bottom half are outgoing connections from each of the 55 ICs, and top half are incoming connections.

Furthermore, we examined the entire distribution of SHAP values, which is shown as a histogram in Figure 6. This demonstrates that all connections contribute to the model’s prediction, albeit most of them to a small degree. The distribution shows considerable spread and a long tail, where the contribution of the most important connection (from IC 34 to IC 24; compare Figure 5) is two orders of magnitude larger than connections at the mode of the histogram.

**Figure 6:**
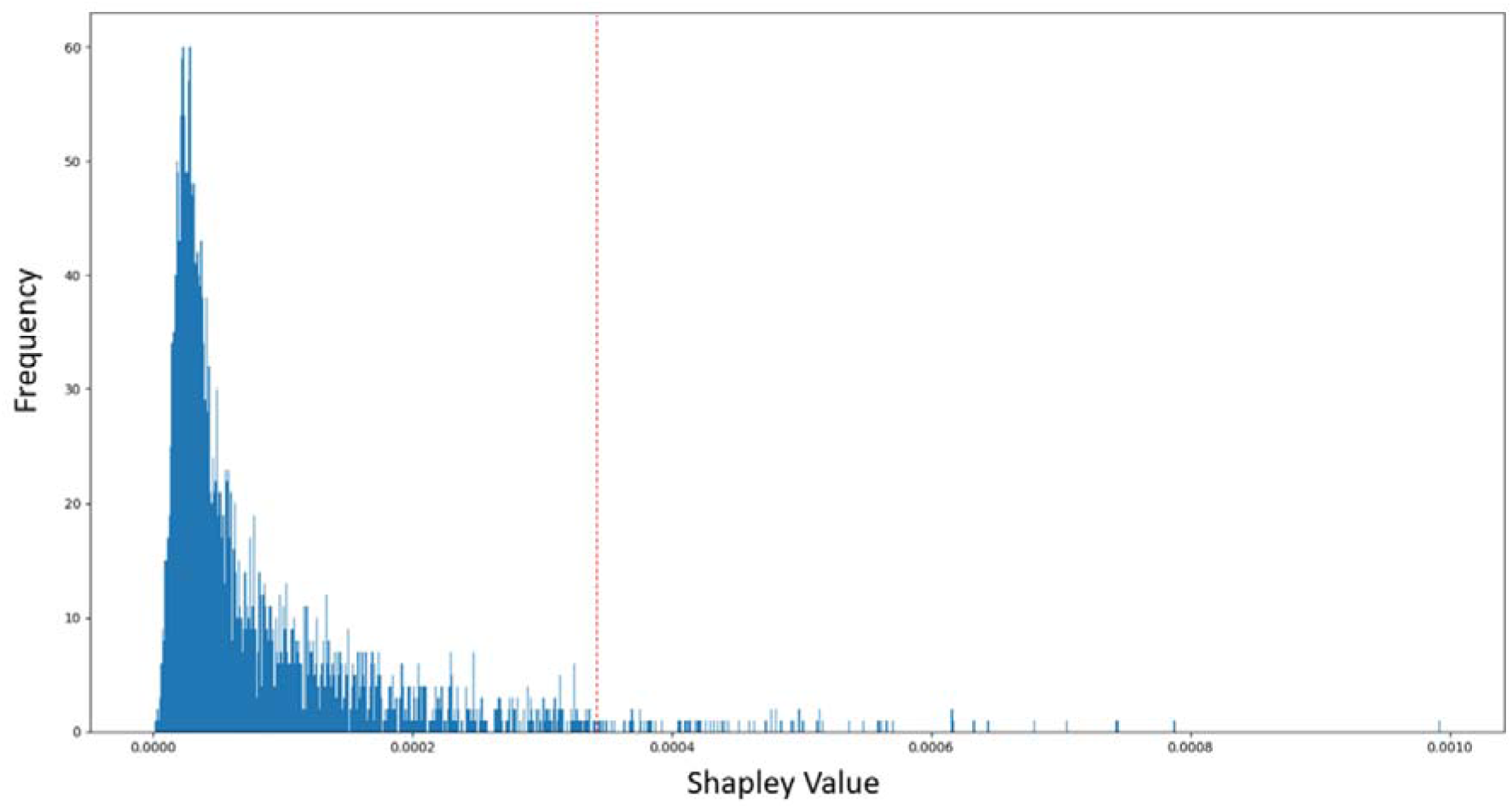
Shapley value distribution for sigmoid SVM paired with rDCM(55) on the test data. Values to the right of the dashed red line are in the top 100.

## Discussion

MDD is a syndrome with heterogenous disease trajectories (Merikangas et al., 1994) and variable treatment responses (Rush et al., 2006). Given the importance for clinical management, predicting future clinical outcomes of individual MDD patients has become an important topic in computational psychiatry. In particular, various fMRI studies have examined the feasibility of predicting treatment response (e.g. Harris et al., 2022; Hopman et al., 2021; Ju et al., 2020; Osuch et al., 2018; Queirazza et al., 2019), relapse (e.g. Berwian et al., 2020; Lawrence et al., 2022), or disease trajectories (e.g. Frässle et al., 2020; Schmaal et al., 2015) in individuals with MDD.

By contrast, there have been hardly any attempts to use fMRI to address another challenge of similar importance: the early detection of individuals who are at risk of experiencing a future episode of depression. Given the high frequency of a prolonged remitting-relapsing disease course after a first episode of MDD (Eaton et al., 2008), identifying at-risk individuals is crucial for enabling the targeted deployment of preventive measures and early interventions. So far, to our knowledge, there has only been a single study that used fMRI for detecting individuals at-risk for future depression (Hirshfeld-Becker et al., 2019). This previous study used rs-fMRI and functional connectivity measures in a small sample of individuals with familial risk for MDD (N=33 for prediction).

The study presented in this paper is novel in several ways. It is the first study using generative models of fMRI data as a basis for predicting future depressive episodes, using three different variants of DCM, in comparison to simpler functional connectivity measures. It uses a large balanced sample size (N=906), carefully matches groups with presence and absence of depressive symptoms, examines the combination of 8 connectivity feature sets with 17 classifiers in a training set, and evaluates the generalisability of the best predictions using a held-out test set.

The results from the training set (Table 1) indicated that the combination of the rDCM(55) feature set (i.e. rDCM-based connectivity estimates between 55 networks or ICs) and a SVM (with a sigmoid kernel) performed best, showing an AUROC of 0.66 and an accuracy of 63%. This result was significantly above chance, as indicated by permutation testing (p=0.001, Figure 3B). Moreover, across classifiers, rDCM demonstrated higher predictive value than other connectivity methods (see Table 1): for 15 out of the 17 classifiers tested, one of the rDCM feature sets resulted in the best AUROC; in 11/15 cases, the best feature set was rDCM(55). Examining the results along the other dimension of our investigation, i.e. across all connectivity feature sets, SVMs performed better than other classifiers: for 6 out of 8 connectivity feature sets, one of the SVM variants had the highest accuracy. In particular, a SVM with a sigmoid kernel performed best for 3 feature sets, surpassing any other classifier.

Evaluating the best combination (i.e. rDCM(55) + SVM with sigmoid kernel) on the test set confirmed the generalisability of the predictions, resulting in an AUROC of 0.64 and an accuracy of 62%. This was significantly above chance (p=0.001), as confirmed by permutation testing (Figure 4B). In a post-hoc analysis, we also evaluated the predictive value of all other connectivity feature sets on the test set; notably, for each feature set, we used the classifier that had performed best on the training set. These analyses showed that three other combinations of connectivity features/classifiers (rDCM(21) + sigmoid SVM, FC(6) + sigmoid SVM, and FC(21) + Gaussian Naïve Bayes) also achieved significant results, although with slightly lower accuracy (58-59%).

In short, our results thus demonstrate that a GE procedure – based on applying rDCM to rs-fMRI timeseries from a large number of ICs (55) – enabled the best predictions about the occurrence of future depressive episodes within a 3-year period. Having said this, the superiority of GE over a simpler prediction procedure based on FC estimates was not large, amounting to 3% higher accuracy and 0.03 higher AUROC compared to the combination of FC(6) + sigmoid SVM. A binomial test indicated that this difference in accuracy was not significant (p=0.315).

The lack of a decisive advantage of generative embedding in this rs-fMRI study contrasts with previous task-based fMRI studies in which GE based on DCM was clearly superior to predictions based on FC estimates (e.g. Brodersen et al., 2011, 2014; Frässle et al., 2018, 2020). For example, DCM estimates of effective connectivity during a face perception task allowed for substantially more accurate predictions of MDD disease trajectories than FC estimates: balanced accuracies for predicting a chronic course vs. remission were 79% for DCM and 50% for FC, a difference that was highly significant (Frässle et al. 2020).

In order to understand the limited advantage of GE over FC-based prediction in this study, it is useful to first consider the general reasons why one would, in general, expect GE to show superior performance. In brief:

i. GE exploits the fact that a generative model partitions data into signal and noise. Using model parameter estimates (as a low-dimensional representation of signal) as features for subsequent ML ensures that only meaningful information underpins training of classifiers. This makes it less likely that predictions are informed by noise and do not generalise. By contrast, measures of functional connectivity, such as correlation coefficients, reflect both signal and noise. As highlighted by Friston (2011), functional connectivity estimates based on correlations are highly susceptible to changes in the signal-to-noise ratio of data.
ii. A generative model like DCM distinguishes different mechanisms how measured signal in a system of interest is caused, e.g. connections between system nodes or external inputs. This allows predictions to be differentially informed by distinct system mechanisms. By contrast, FC cannot distinguish whether co-varying signal in two brain regions is caused by shared input or by connections between the regions.
iii. DCM provides directed connectivity estimates, allowing one to obtain separate weights for reciprocal connections between regions. By contrast, FC can only provide undirected estimates of connection strengths.
iv. From a classical test theory perspective, test-retest reliability of connection strength estimates would be considered an important prerequisite for predictive validity. Concerning rs-FC, test-retest reliability has been examined in numerous studies; a recent meta-analysis reported that, on average, individual connection estimates have limited test-retest reliability (Noble et al., 2019). A direct comparison between FC and rDCM-based estimates of connectivity on identical data (rs-fMRI and multiple tasks) demonstrated that rDCM performed more favourably in this regard (see Figure 3 in Frässle & Stephan, 2022).

Considering these general factors, one possibility why we only found a limited advantage of GE over FC-based predictions in this study relates to (i) above: in the present study, connectivity was estimated from timeseries that resulted from ICA decomposition and subsequent (manual) removal of components that were identified as noise (Alfaro-Almagro et al., 2018). This approach may have diminished the difference between GE and FC-based prediction with regard to denoising. For comparison, in previous comparisons of GE and FC-based predictions (e.g. Brodersen et al., 2011, 2014; Frässle et al., 2018, 2020), timeseries were obtained by computing the first principal component from regional BOLD measurements, which does not involve a specific distinction between signal and noise. Another possible explanation derives from (ii): application of rDCM to rs-fMRI data essentially means that the model “switches off” external inputs (Frässle, Harrison, et al., 2021). This reduces the superiority in representational richness of GE.

In summary, this suggests that, in the current setting of IC-based rs-fMRI timeseries, only factors (iii) and (iv) – but not factors (i) and (ii) – could potentially contribute to higher performance of GE. In order to obtain an impression of the potential impact of factor (iii) – the ability of DCM to obtain separate weights for reciprocal connections between network nodes – we visually explored the asymmetries of node-level SHAP values for incoming versus outgoing connections. For each of the 55 network nodes (ICs), Fig. 7 plots SHAP values summed across all incoming (afferent) and outgoing (efferent) connections, respectively. Visually, it is apparent that for many of the network nodes, the explanatory contributions of incoming versus outgoing connections differ considerably (up to 59%). A more fine-grained plot of connection-specific SHAP values is provided by Fig. 8.

**Figure 7:**
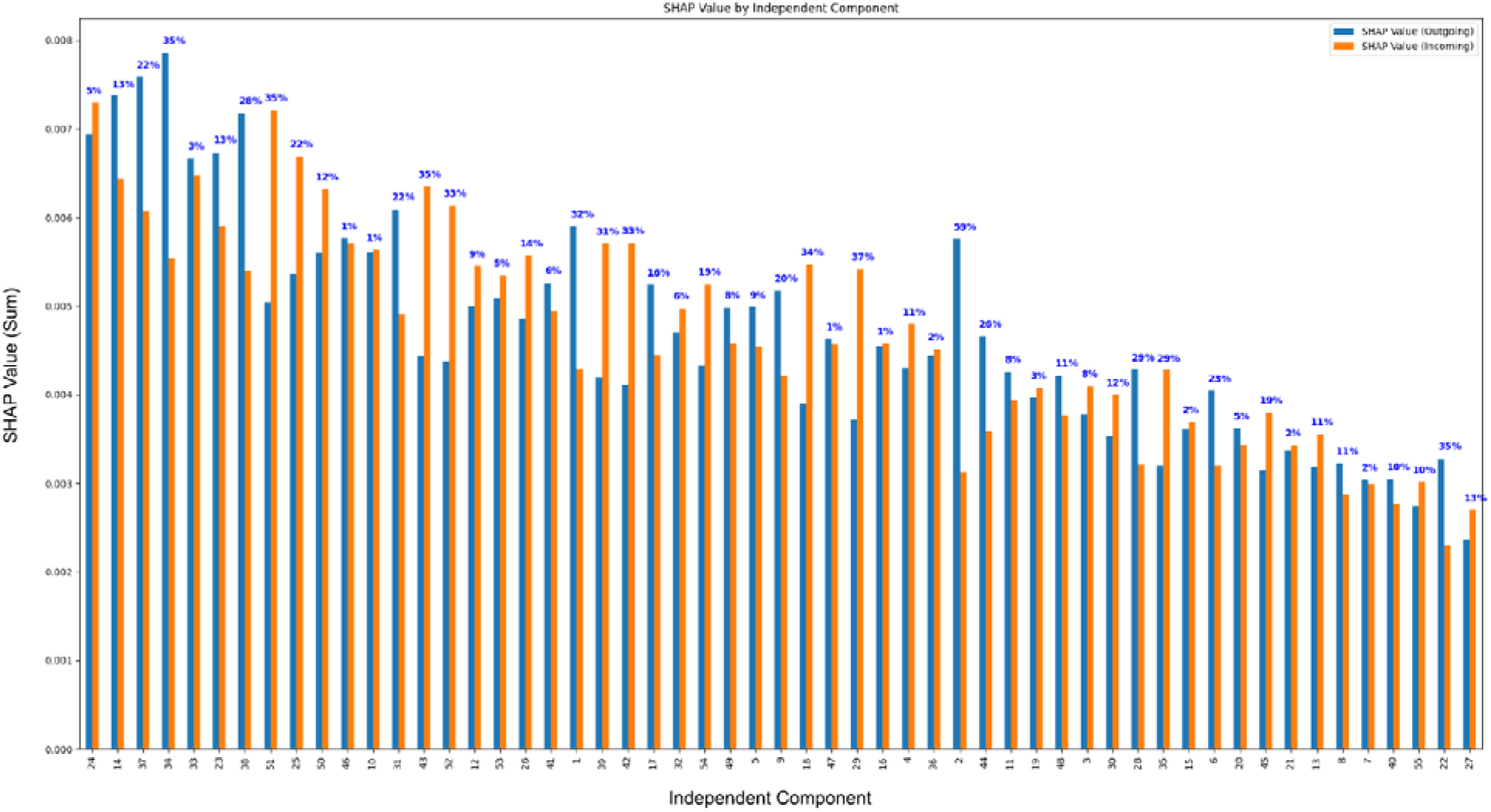
SHAP values summed across the incoming (afferent) and outgoing (efferent) connections of each IC. Percentages indicate the % difference in SHAP values for afferent and efferent connections. The plot concerns predictions based on rDCM(55) estimates and SVM with a sigmoid kernel.

**Figure 8:**
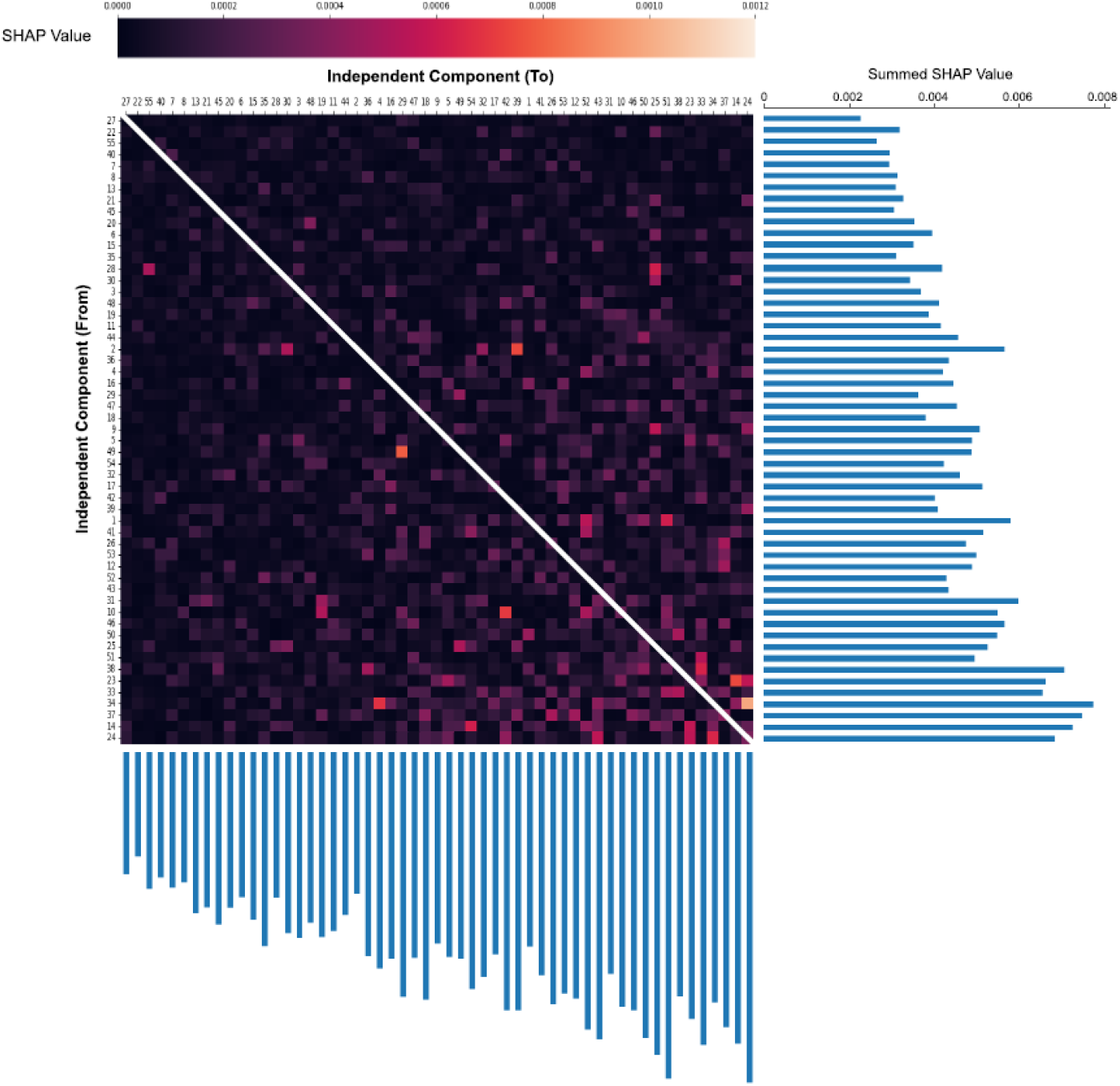
Matrix of connections between all ICs, showing the connections’ colour-coded SHAP values (test data) for predictions based on rDCM(55) estimates and sigmoid SVM. ICs are ordered according to summed SHAP values.

These plots also illustrate a disadvantage of the analysis approach we have chosen in the current study. Specifically, using ICs as network nodes diminish the advantage GE usually enjoys in terms of rendering predictions neurophysiologically interpretable. For example, as shown by Figures 5 and 8, the connection with the largest SHAP value is the connection from IC 34 to IC 24. Both of these components include a set of fronto-parietal areas: IC 34 includes bilateral frontal regions that appear to match the location of the frontal eye fields as well as more anterior parts of the superior parietal cortex. By contrast, IC 24 contains more posterior bilateral parietal areas, including large parts of bilateral intraparietal sulcus, as well as parts of right middle frontal gyrus and right middle/inferior temporal gyrus. Given this complex anatomical configuration, the biological interpretation of a (directed) functional coupling between IC 34 and IC 24 is not as straightforward as a functional coupling between specific frontal and/or parietal areas. While the FC between these components does not enable any easier interpretations, this example illustrates that the usual interpretive advantage of GE tends to be lost when using IC components as nodes of networks.

Three further aspects of the results deserve discussion. First, it may initially seem surprising that SVM turned out to be the most successful classifier in our comparison, surpassing potentially more powerful methods like neural networks. However, this result is compatible with several recent reports that, for neuroimaging data, kernel-based methods like SVMs (and, in some cases, even simpler linear models) perform equivalently to neural networks for sample sizes up to 10,000 (Cole et al., 2017; He et al., 2020; Schulz et al., 2020).

Second, in our post-hoc analysis of connectivity features/classifier combinations on the test set, three of four significant predictions used the same classifier, an SVM with a sigmoid kernel. Strikingly, FC achieved a significant 59% predictive accuracy using only 6 ICs, whereas the more accurate prediction by rDCM (62%) used 55 ICs, respectively. The resulting difference in the number of features is substantial (15 for FC versus 3025 for rDCM), and it is not immediately clear why predictions based on functional vs. effective connectivity differed greatly in the preferred dimensionality of the feature set. One speculative explanation – which would be consistent with the findings in Figures 7 and 8 – is that differences in the strengths of reciprocal between-network connections provide subtle but meaningful information that is distributed over many connections (compare factor (iii) above). This type of information would only be reflected by rDCM, but not by FC-based, connectivity estimates. More generally, it is not clear why FC(6) performed so well on the test data at all. The nested CV on the training data did not indicate that this feature set might be particularly predictive (maximum accuracy of FC-based predictions with any classifier was 54%, none of them significant). The finding of a higher accuracy (59%) on the test set our post-hoc analysis was surprising. It might be a chance result due to the variance inherent in CV procedures (Varoquaux, 2018) but otherwise lacks a compelling explanation.

Third, contrary to our expectations, predictions based on stochastic and spectral DCM did not generalise to the test set. One possible reason for the lack of successful generalisation is that the higher complexity of the model formulation (e.g. the flexible hemodynamic component and the more sophisticated noise model) could make parameter estimation less reliable, e.g. due to greater abundance of local extrema in the objective function, which would be expected to harm generalisability. This possibility is supported by a recent investigation of parameter recovery of spectral DCM and rDCM which found more accurate parameter recovery for the latter (Frässle et al. 2021). Perhaps even more importantly, however, we could only run spectral and stochastic DCM for 6 ICs on our cluster; for larger feature sets, their compute time (within the context of our entire analysis pipeline) became prohibitively long. However, considering the success of rDCM based on 55 ICs, it is plausible that spectral and stochastic DCM may have performed better if we had been able to run them with larger IC sets (21 and 55).

How does the prediction performance achieved in this study compare to previous results in the literature? The only previous fMRI study on predicting future depression (Hirshfeld-Becker et al., 2019) used FC estimates based on rs-fMRI data from six regions, achieving 92% accuracy. However, this study recruited never-depressed children with familial risk for MDD, as opposed to never-depressed participants from the general population as in our study. Additionally, given the more specific focus of the previous study, only 33 participants were available for classification (25 at-risk children, eight controls); this small sample size did not allow for verification in a held-out dataset. Another useful (although not fMRI-based) comparison study utilized structural MRI together with clinical data, questionnaires, and environmental variables (Toenders et al. 2021). The study used a large training set (N=407 adolescents) and an independent test set (N=137), achieving an AUROC between 0.68-0.72.

It is also instructive to consider the results from non-imaging studies that used demographic, socioeconomic, and clinical variables for predicting the future onset of depression. When considering those studies that had large sample sizes (i.e. N>500) and tested for generalisability in an independent test set, the reported AUROC values in the literature range between 0.71-0.87 (Caldirola et al., 2022; King et al., 2008; Librenza-Garcia et al., 2021; Na et al., 2020; Xu et al., 2019). It is noteworthy, however, that these studies mostly used imbalanced datasets where the number of negative cases (no future depressive episode) far outnumber the positive cases. For example, in the two studies with the highest prediction performance – i.e., AUROC of 0.87 (Na et al., 2020) and 0.85 (Caldirola et al., 2022) – individuals with future depressive episodes amounted to approx. only 8% and 7% of the respective samples. Even when techniques such as oversampling are used (as in Na et al., 2020; but not always the case in other studies), such imbalance can lead to overly optimistic estimates of prediction performance.

Our study has strengths and limitations. Its strengths include an ex ante analysis plan (https://gitlab.ethz.ch/tnu/analysis-plans/galioullineetal_ukbb_pred_depr) and a large (N>900) and balanced sample in which groups were carefully matched for 7 potentially confounding variables (age, sex, handedness, tobacco smoking frequency, alcohol consumption frequency, ongoing addictions to illicit drugs, and historical cannabis consumption). This degree of matching is unusually comprehensive (for comparison, in clinical trials and observational studies, it is rarely possible to match for more than two variables) and only made possible by the large resource of the UK Biobank. Furthermore, we conducted a comprehensive comparison of 8 different connectivity measures and 17 classifiers, ensuring that training and test data were strictly separated throughout all analyses.

Concerning weaknesses, our study has a retrospective design which allows for less robust conclusions than from a prospective study. Furthermore, one potential weakness of the variants of DCM used in this study is that they all rely on variational Bayesian techniques, rendering model inversion susceptible to local extrema in the objective function (Daunizeau et al., 2011). In theory, this could have been addressed by a multi-start procedure, as in previous work with DCM (Schöbi et al., 2021; van Wijk et al., 2018). In practice, however, we were unable to implement this approach given that it would have led to an explosion of the already very substantial compute time. Finally, the greatest limitation of our study is the definition of depressive episodes. Given the heterogeneity of clinical data in the UK Biobank and the lack of systematic information about absence/presence of a clinical diagnosis of depression, we combined multiple sources of information within UK Biobank – i.e., clinical records, questionnaires (PHQ, MHQ) and self-report specifically on issues of depression – to identify indicators of at least one depressive episode within three years after the fMRI scan. Clearly, this partial reliance on self-report is not ideal; additionally, the resulting group of participants with a putative depressive episode (D+ group) is likely heterogeneous and might include people with very different severities of depression. Furthermore, there is rarely information on when exactly within the 3-year period a depressive episode occurred; the likely interindividual variability in the latency of symptom onset after the fMRI scan would further add to the heterogeneity of the D+ group. Having said this, our approach is similar to previous analyses of depression in the UK Biobank that also relied on self-report and questionnaires like the MHQ (Howard et al., 2020). More generally, a pragmatic approach to identifying individuals with likely clinical characteristics is often unavoidable when working with large heterogeneous databases (for an example using self-reported depression in genetics, see Wray et al., 2018). The challenge how to optimally extract data from the UK Biobank for studies of MDD is being addressed by ongoing methodological developments (Dutt et al., 2022) which will help to improve and standardise future studies.

Overall, our results have four implications. First, given the challenging nature of the prediction problem tackled in the study (i.e. occurrence of indicators of depressive episodes, as opposed to full clinical diagnoses, over a three year period), it is encouraging that significant predictions on held-out data can be obtained at all. Second, despite this success and the potential for further optimisation, our study suggests that fMRI on its own may not be sufficient for clinically useful predictions. Future studies of predicting depression should utilise fMRI-based connectivity estimates in conjunction with additional data (e.g. demographic, socioeconomic, clinical). Third, while GE results based on rDCM were consistently successful across all classifiers and enjoyed a numerical advantage over FC for clinical predictions, performance differences were modest and nonsignificant. The magnitude of performance differences between GE and FC in this study and previous work suggests that adding task-based fMRI may enhance the difference in predictive accuracy. Finally, using IC components as network nodes diminishes the usual advantage of GE with regard to biological interpretability of predictions. In order to maintain the interpretability of GE based predictions, it would seem advantageous to compute effective connectivity between disjoint areas from parcellations based on combined anatomical-functional criteria (e.g. Fan et al., 2016; Glasser et al., 2016). We hope that these conclusions will be useful for future work on predicting the occurrence of depressive episodes.

## Data Availability

All data produced in the present study are available upon reasonable request to the authors

## Author contributions according to CRediT (Contributor Roles Taxonomy)

**Herman Galioulline:** Investigation, Software, Formal analysis, Writing – Original Draft, Visualization

**Stefan Frässle:** Investigation, Supervision, Validation, Software, Methodology, Writing – Review & Editing

**Samuel Harrison:** Investigation, Supervision, Validation, Software, Methodology, Writing – Review & Editing

**Inês Pereira:** Investigation, Validation, Writing – Review & Editing

**Jakob Heinzle:** Investigation, Supervision, Validation, Software, Methodology, Writing – Review & Editing

**Klaas Enno Stephan:** Investigation, Conceptualization, Supervision, Methodology, Project administration, Funding acquisition, Writing – Review & Editing

## Acknowledgements

This work was supported by the René and Susanne Braginsky Foundation (KES), the ETH Foundation (KES), and project grant 320030_179377 by the Swiss National Science Foundation (KES).

